# Safety and Feasibility of a “Fast-Track” Monitoring Protocol for Patients Treated with Intravenous Thrombolytic Therapy

**DOI:** 10.1101/2023.03.27.23287831

**Authors:** Keiko A Fukuda, Kavit Shah, Cynthia Kenmuir, Derrick Barnagian, Baraa Nawash, Mackenzie Nelson, Shashvat Desai, Marcelo Rocha, Matthew Starr, Eileen Roach, Stephanie Henry, Bradley J Molyneaux, Ashutosh P Jadhav

## Abstract

**Introduction:** Our health care systems continue to face significant strain due to chronically taxed intensive care resources. A subgroup of post-thrombolytic stroke patients may not require prolonged intensive monitoring, alleviating some burden. Here we describe the safety, feasibility, and utility of a Fast-Track Protocol (FTP) for early de-escalation of high-acuity monitoring.

**Methods:** We compared a prospective cohort of FTP patients at our stroke centers from April 2020 – February 2022 to a similar retrospective cohort. Those on presentation with NIHSS < 10 and without large vessel occlusion or flow-limiting stenosis, intravenous anti-hypertensive use, and any hemodynamic or respiratory concerns were eligible. Primary outcomes included early neurologic deterioration, defined as worsening of NIHSS ≥ 4-points at 24 hours, parenchymal hemorrhage, and symptomatic intracranial hemorrhage.

**Results:** Of 574 thrombolysis patients, 119 (21%) were eligible for FTP. One hundred (88%) were included for analysis. The median ± IQR hospitalization was 2.0 ± 1.6 days. None of the four patients with early neurologic deterioration were due to hemorrhage. No symptomatic intracranial hemorrhages occurred, and no FTP patients were transferred back to the ICU. Median ± IQR 90-day modified Rankin scale was 1 ± 1.

**Discussion:** FTP is a safe and feasible strategy to triage ICU patients and decrease unnecessary ICU monitoring. This is important in a post-pandemic era as ICU resources continue to fluctuate. Future studies are needed to establish the optimal level of monitoring in post thrombolytic patients.

## Introduction

Patients who receive intravenous thrombolysis (IVT), irrespective of endovascular thrombectomy (EVT), are commonly monitored in a high acuity setting, such as an intensive care unit (ICU) or dedicated stroke unit, for a minimum of 24 hours with frequent assessments of vital signs and neurological exams. [1] This ensures these patients receive close observation to detect any change in their neurologic condition, which would warrant emergent medical or surgical intervention. More recently, these ICU level resources have posed greater challenges for health care systems.

The Coronavirus disease-2019 (COVID-19) pandemic changed the healthcare landscape, especially in critical care settings, due to the rapid rise in patient volumes, with nearly 100 million Americans suffering infection and over 1 million deaths to date. [2, 3] Patients suffering acute ischemic strokes (AIS) invariably require evaluation and rapid treatment with IVT or EVT if indicated, with subsequent admission to the hospital for intensive care, diagnostic investigation, and management. Health systems of care must adapt to the increased use of interventions for stroke treatment and the risk of future waves of infection, which continue to threaten intensive care resources.

Those who experience an NIH Stroke Score (NIHSS) decline of ≥ 4 points within 24 hours of IVT are defined as having early neurological decline (END). [4] Furthermore, if the etiology of END is secondary to an intracranial hemorrhage (ICH), this is defined as a symptomatic intracranial hemorrhage (sICH). [5] This feared complication reassuringly occurs in a minority of patients. [6, 7] An earlier retrospective database analysis found a mean time from intravenous tissue plasminogen activator (IV tPA or IV alteplase) administration to END of less than 5 hours, with over 80% of cases occurring within 12 hours. [8] Several factors were predictive of END and any ICH, including an untreated large vessel occlusion and higher baseline NIHSS, consistent with prior studies. [9-11] Moreover, the OPTIMIST safety trial found all patients in their population of NIHSS less than 10 on presentation without initial ICU needs remained clinically stable and could be safely monitored with a lower intensity protocol. [12] Given these findings and chronically taxed ICUs, we implemented a Fast-Track Protocol (FTP) to permit early de-escalation of high acuity clinical monitoring in a selected cohort of IVT patients. We sought to evaluate the protocol’s safety, feasibility, and utility on our selected patient population.

## Methods

After Quality Improvement Review Committee approval, we performed a retrospective analysis of a prospectively maintained database of AIS receiving IVT (either intravenous Alteplase or Tenecteplase) between April 2020 through February 2022. From April 2020 – March 2021, patients received IV alteplase and after March 24, 2021, all patients received IV Tenecteplase consistent with our health systems’ planned change, unrelated to our study. Demographic characteristics, treatment information, and clinical and radiological data were extracted and compared to a similar retrospective database of AIS patients at our institution who received IV Alteplase from 2013-2019. [8]

### Patient Selection

All AIS patients who received IVT after presenting directly or transferring to one of our comprehensive stroke centers (CSC) or thrombectomy-capable stroke center (TSC) from one of our referring hospitals during the study period were included for analysis. These patients must have presented within 4.5 hours of last seen well (LSW) and received IVT based upon AHA/ASA guidelines. All patients were subsequently admitted to the ICU for post-IVT monitoring and management. In addition, patients were eligible for and enrolled in FTP based on the following criteria: presenting NIHSS < 10, no intravenous anti-hypertensive medication administered to maintain blood pressure under established guidelines of 180/105; absence of large vessel occlusion (LVO) on non-invasive angiographic imaging, moderate to severe extracranial internal carotid artery (ICA) stenosis, or intracranial ICA stenosis; and no hemodynamic or respiratory concerns. For patients to remain in the FTP, they must have maintained an NIHSS of less than 10, not received any intravenous anti-hypertensive medication to maintain blood pressure under established guidelines and have no additional hemodynamic or respiratory concerns during their ICU stay. Before transferring out of the ICU, a vascular neurology fellow or neurocritical care or vascular neurology attending must have reviewed a 12-hour CTH or MRI to ensure there was no evidence of hemorrhagic conversion or finding warranting a continued higher level of care (Figure 1a). Patients in the historical cohort were previously described, [8] and were monitored by AHA Guidelines (vital signs and neurologic checks every 15 minutes for 2 hours, every 30 minutes for 6 hours, and every hour for 16 hours) (Figure 1b). [13]

**Figure 1a.**
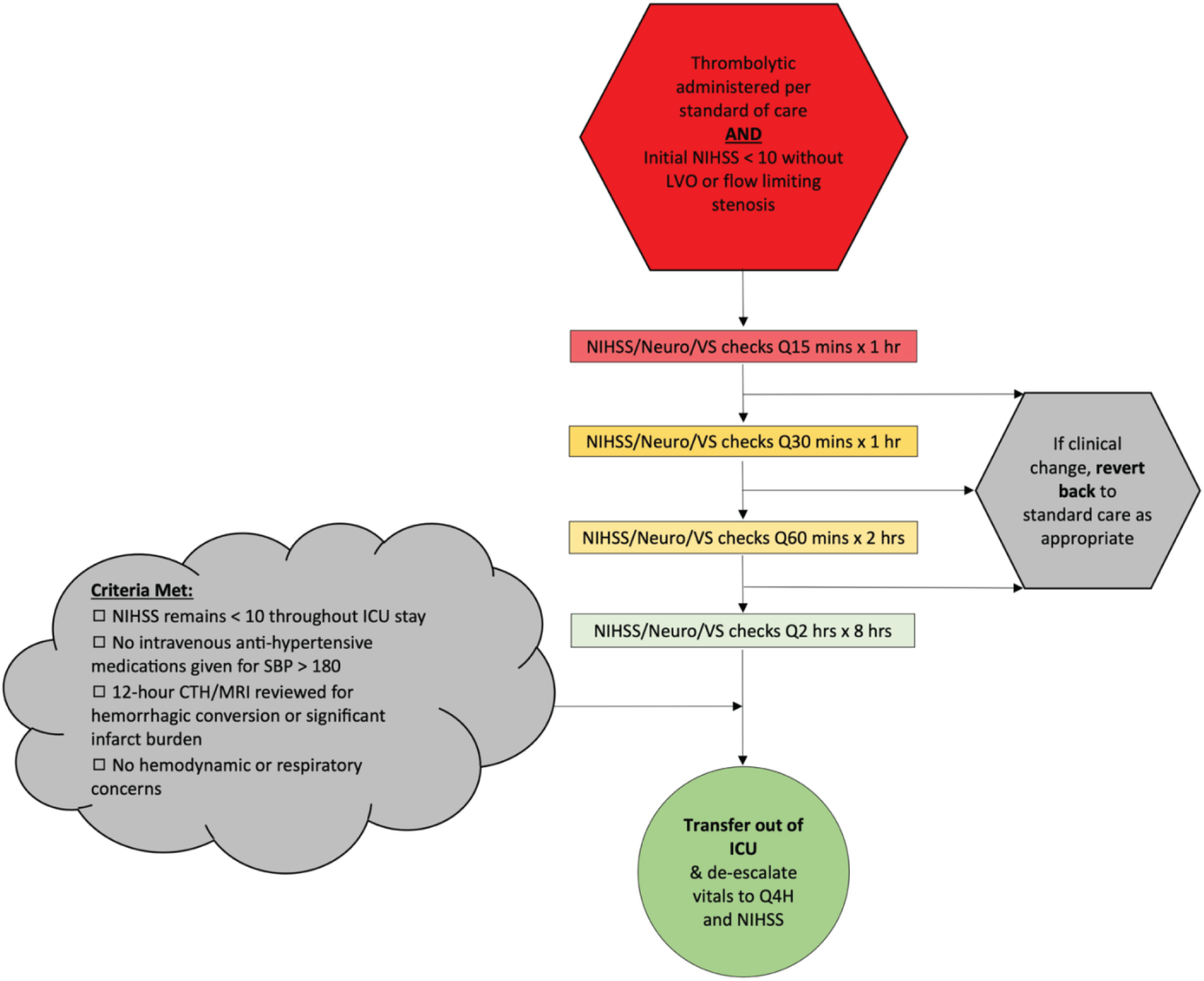
Fast-Track Protocol.

**Figure 1b.**
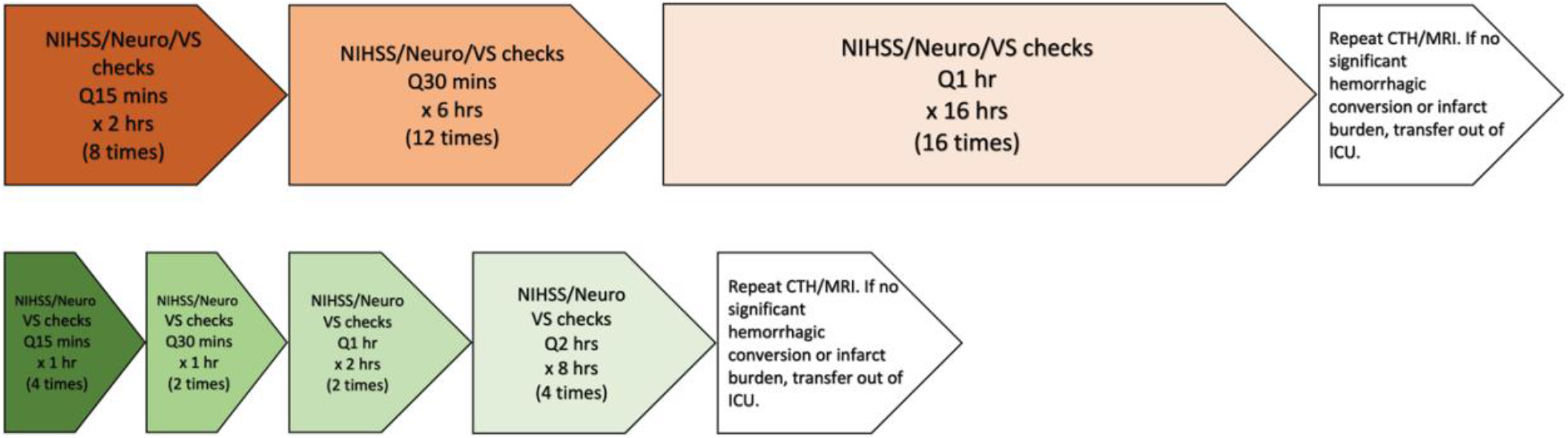
Standard Monitoring Protocol versus Fast-Track Protocol.

### Baseline Characteristics

Baseline demographic (age, sex, race), clinical (presenting NIHSS, NIHSS at all pre-specified intervals within 24 hours of IVT administration, LSW, risk factor profile, baseline modified Rankin scale [mRS]), and radiographic (pre-treatment ASPECTS, presence of LVO, and post-IVT CTH findings) information were collected and analyzed by a vascular neurologist (KS, BJM, KAF) blinded to patient characteristics.

### Outcomes

Primary outcomes included END (≥ 4-point NIHSS worsening at 24 hours) from any cause, parenchymal hemorrhage (PH1 or PH2), and sICH. [5]

### Statistical Analyses

Continuous variables are reported as mean ± SD or median with interquartile range (as appropriate), and categorical variables are reported as proportions. Between-groups comparison for continuous variables was performed using Student’s t-test, multiple t-test or Mann-Whitney U test, as appropriate, and categorical variables using the chi-squared test or Fisher exact test, as appropriate. Significance was defined as p ≤ 0.05. Images were developed using Microsoft Office version 2022. Statistical analysis was performed using Stata version MP 14.0 (StataCorp, College Station, TX).

## Results

A total of 574 patients received IVT from April 2020 through February 2022. Seventy-nine percent were not eligible for FTP as they did not meet criteria with an NIHSS on presentation of at least 10, an LVO or flow-limiting stenosis, or use of IV anti-hypertensive medications. Of those who received IVT, 119 (21%) were intended for FTP. One hundred (17%) of these patients were included for analysis. Of the excluded patients, 14 had FTP criteria violations on admission upon further review, three were eligible and intended for FTP but not timely protocolized, and two reverted to the standard protocol due to hemodynamic instability (Figure 2). Mean age was 61 ± 14 years, 48% were women, and mean NIHSS on presentation was 4.4 ± 2.4, which was significantly lower than the retrospective population. Remaining baseline characteristics are as shown in Table 1, without significant differences between the two populations.

**Table 1.** Baseline Patient Characteristics

|  | **Characteristics** | Fast Track pts<br>n=100 | 2013-2019<br>FTP eligible<br>pts<br>n=315 |
| --- | --- | --- | --- |
| Demographics | Age, mean ± SD | 61 ± 14 | 64 ± 15 |
|  | Female, n (%) | 48 (48) | 147 (47) |
| | Mean NIHSS Pre-IVT ± SD | 4.4 ± 2.4 | 5.5 ± 2.0 |
| Risk factors | Hypertension, n (%) | 61 (61) | 207 (66) |
|  | Diabetes Mellitus, n (%) | 26 (26) | 95 (30) |
|  | Hyperlipidemia, n (%) | 52 (52) | 159 (50) |
|  | Smoking History, n (%) | 23 (23) | 83 (26) |
|  | Atrial fibrillation, n (%) | 5 (5.6) | 235 (19) |
| Hospital admission | Presented to CSC/TSC ED, n (%) | 42 (42) | 90 (29) |
|  | Median Lytic to Fast Track (h:m) | 1:56 |  |
| Diagnosis | Stroke, n (%) | 55 (55) |  |
|  | Seizure, n (%) | 4 (4) |  |
|  | Stroke Mimic (migraine, conversion disorder), n (%) | 31 (31) |  |
|  | Other (toxic/metabolic, AMS, peripheral), n (%) | 10 (10) |  |

**Figure 2.**
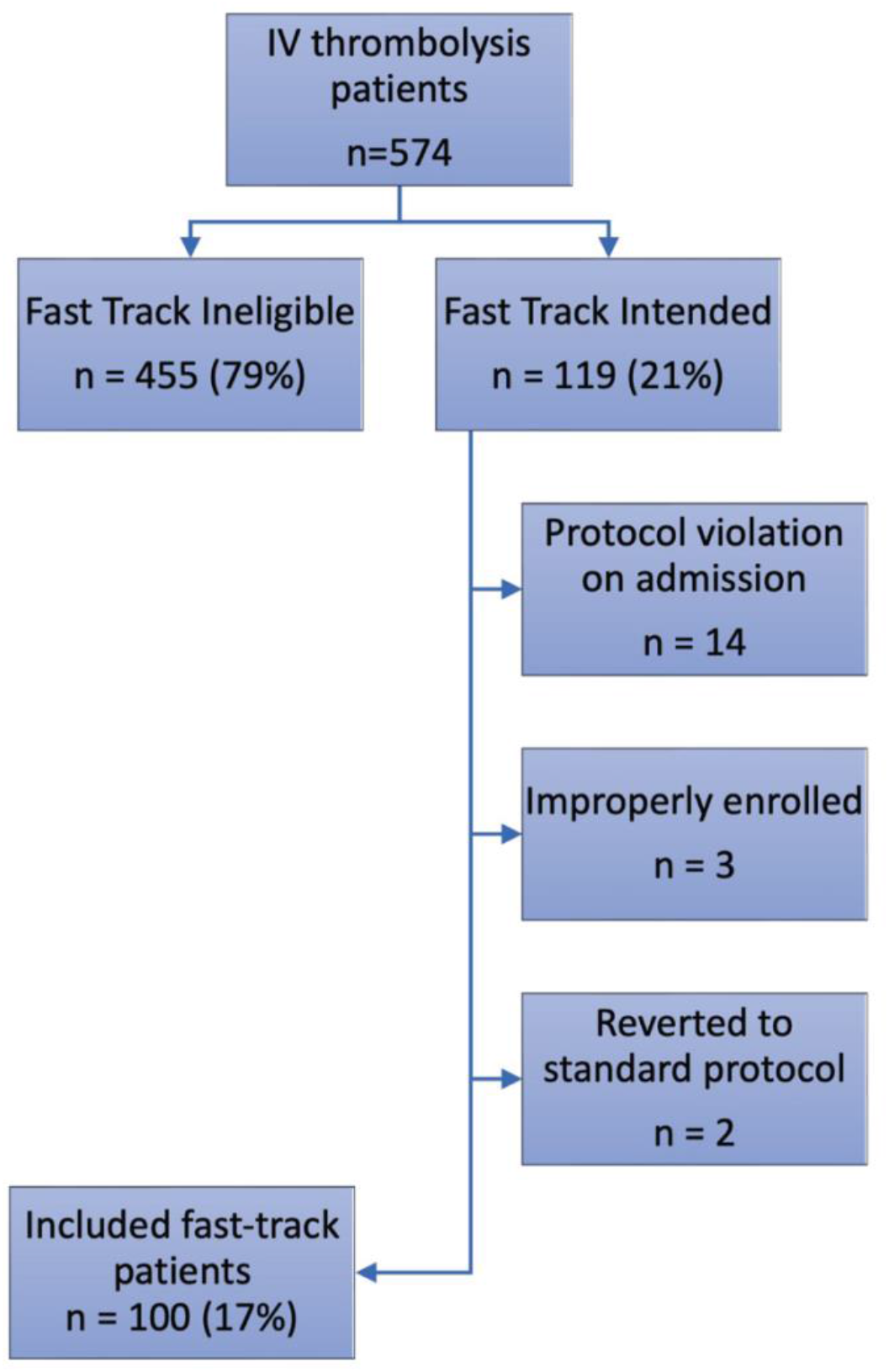
Flow chart of excluded and included patients.

Amongst the FTP population, 42% presented directly to our CSC or TSC hospitals, which was significantly higher than our retrospective cohort (Table 1). This was likely due to the inclusion of our rural TSC in our current cohort. Median time from IVT to initiation of FTP was 1 hour and 56 minutes. Only 55% had a final diagnosis of stroke. Of the remaining, 31% had a stroke mimic (migraine or conversion disorder), 10% had alternate etiologies for neurologic deficits (encephalopathy, metabolic/toxic causes, or peripheral nerve disorder), and 4% were secondary to seizure. Median ± IQR length of hospitalization was 2.0 ± 1.6 days. Discharge disposition was largely to home (74%), followed by inpatient rehabilitation (23%) and skilled nursing facility (3%). No patients expired in the hospital in the FTP group.

Regarding our primary outcomes, no symptomatic intracranial hemorrhages occurred, and none of the FTP patients required transfer back to the ICU or escalation of care due to clinical deterioration after transfer out. Four patients experienced END, defined as an increase in NIHSS 4 or more points from presentation within the first 24 hours. Three cases of END had NIHSS difference of 4 or more immediately upon transfer to our CSC or from ED to ICU, likely representing inter-institutional or interrater variability. One case was due to symptomatic worsening of stroke mimics, including conversion and migraine disorder without acute infarcts found on MRI. One patient reverted to the standard post-thrombolytic protocol due to infarct evolution without hemorrhagic transformation. None were due to intracranial hemorrhage for any reason. Median ± IQR 90-day modified Rankin score was 1 ± 1. There was no significant difference in END, ICH, or median hospitalization length compared to a historical cohort that received standard monitoring, including hourly neurological exams, for the full 24 hours (Table 2).

**Table 2.** Safety outcomes

| **Safety Outcomes** | Fast Track<br>n=100 | 2013-2019 FTP<br>eligible pts<br>n=315 | p-<br>value |
| --- | --- | --- | --- |
| Home, n (%) | 75 (75) | 233 (74) | 0.83 |
| Inpatient Rehab, n (%) | 22 (22) | 67 (31) |  |
| SNF, n (%) | 3 (3) | 13 (4) |  |
| Expired, n (%) | 0 (0) | 2 (0.6) |  |
| END, n (%) | 4 (4) | 9 (3) | 0.56 |
| sICH, n (%) | 0 (0) | 1 (0.3) | 0.57 |
| Median Hospitalization Length,<br>d (IQR) | 2.0 ± 1.6 | 2.2 ± 2.1 | 0.78 |

## Discussion

FTP demonstrates a safe method of transferring post-thrombolytic patients out of the ICU who are unlikely to deteriorate and require additional monitoring. None of our patients experienced a primary outcome of intracranial hemorrhage for any reason, most importantly sICH. Selecting for patients with low initial NIHSS without flow-limiting stenoses, large vessel occlusions, or hemodynamic changes likely contributed to the protocol’s overall safety as well as high rate of stroke mimics (31%). The cases in which END occurred were low-risk and did not pose significant safety concerns inherent to FTP. The one patient with continued evolution of their infarct was appropriately identified and reverted to the standard monitoring protocol without incident.

Previous studies of stroke systems of care show that over 80% of those with END occurs within the first 6 hours from IVT, providing further proof of the safety of transferring patients out of the ICU sooner. [8] The guidelines to ensure adequate care of neuro-critically ill patients during a pandemic highlights the need for alternate strategies beyond currently routine practices. [14, 15]

Potential solutions to acute shortages include increasing ICU capacity by discharging suitable patients earlier to lower intensity units. The unique nature of the COVID-19 pandemic has led us to carefully develop new guidelines for triage with such strategies as the FTP to ensure ICU and Stroke Unit resources are reserved for those who will benefit the most and survive with a good quality of life. [3, 14, 15]

Despite a reduction in hospitalizations and utilization of critical care services [16, 17], there was no significant increase in mortality after the onset of the COVID-19 pandemic among critically ill neurologic patients that needed ICU care. [18] This may indicate that ICU care was likely not impacted at most centers. Previous data has shown that among patients hospitalized with these diagnoses but not receiving critical care, there was a small increase in mortality during the pandemic compared to previous years, possibly due to reduced ICU bed availability leading to patients that would ordinarily have been cared for in the ICU being managed in non-critical care settings and by providers that lack specific expertise. [19] The FTP allows for greater bed availability for the critically ill. While not significant, FTP did show a trend toward shortening the overall length of stay, which could assist in decompressing hospitals and reduce associated hospitalization costs.

Strengths of this study include its prospective nature to provide stronger support for earlier deescalation of monitoring in these lower risk patients. However, there was a notable amount of excluded protocol violations on enrollment, primarily due to NIHSS of greater than 9 on presentation. Other violations included the presence of a flow-limiting stenosis and the use of IV anti-hypertensive medications before transfer. Of note, no instances of END occurred in those excluded due to protocol violations. Such enrollment errors should be considered and expected when new protocols such as FTP are implemented into long-standing institutional practices, even at experienced centers like ours. The majority of subjects were also transferred from referring facilities after receiving IVT. This led to a longer median period from IVT to FTP initiation and subsequently may have resulted in the possibility of excluding a subset of patients who became symptomatic prior to initiating FTP in our cohort.

## Conclusion

FTP for selected post-IVT stroke patients is a safe and feasible strategy to triage ICU patients for transfer and decrease unnecessarily prolonged ICU monitoring. This strategy is particularly relevant in a postpandemic era as the need for ICU beds will continue to fluctuate for the foreseeable future. Future randomized controlled studies are necessary to further clarify the optimal level of monitoring in post thrombolytic patients.

## Data Availability

Data used to prepare this manuscript may be made available upon reasonable request.

## Acknowledgements

A.P.J and B.J.M contributed to the conception and design of the study. K.A.F., K.S., C.K., D.B., B.N., M.N., S.D., M.R., M.S., E.R., and S.H. contributed to the acquisition and analysis of the data. K.A.F., K.S., B.J.M, and A.P.J. contributed to drafting of the text and preparation of the figures.

## Abbreviations

(FTP): Fast Track Protocol
(IVT): Intravenous Thrombolysis
(EVT): Endovascular Thrombectomy
(END): Early Neurological Decline
(LSW): Last Seen Well

